# Whole Genome DNA and RNA Sequencing of Whole Blood Elucidates the Genetic Architecture of Gene Expression Underlying a Wide Range of Diseases

**DOI:** 10.1101/2022.04.13.22273841

**Authors:** Chunyu Liu, Roby Joehanes, Jiantao Ma, Yuxuan Wang, Xianbang Sun, Amena Keshawarz, Meera Sooda, Tianxiao Huan, Shih-Jen Hwang, Helena Bui, Brandon Tejada, Peter J. Munson, Demirkale Cumhur, Nancy L. Heard-Costa, Achilleas N Pitsillides, Gina M. Peloso, Michael Feolo, Nataliya Sharopova, Ramachandran S. Vasan, Daniel Levy

**Affiliations:** Department of Biostatistics, School of Public Health, Boston University, Boston, MA, USA; Framingham Heart Study, Framingham, MA, USA; Population Sciences Branch, Division of Intramural Research, National Heart, Lung, and Blood Institute, National Institutes of Health, Bethesda, MD, USA; Nutrition Epidemiology and Data Science, Friedman School of Nutrition Science and Policy, Tufts University, Boston, MA, USA; University of Massachusetts Medical School, Worcester, MA, USA; Departments of Medicine and Epidemiology, Boston University Schools of Medicine and Public Health, Boston, MA, USA

## Abstract

To create a scientific resource of expression quantitative trail loci (eQTL), we conducted a genome-wide association study (GWAS) using genotypes obtained from whole genome sequencing (WGS) of DNA and gene expression levels from RNA sequencing (RNA-seq) of whole blood in 2622 participants in Framingham Heart Study. We identified 6,778,286 *cis*-eQTL variant-gene transcript (eGene) pairs at *p*<5×10^−8^ (2,855,111 unique *cis*-eQTL variants and 15,982 unique eGenes) and 1,469,754 *trans*-eQTL variant-eGene pairs at *p*<1e-12 (526,056 unique *trans*-eQTL variants and 7,233 unique eGenes). In addition, 442,379 *cis*-eQTL variants were associated with expression of 1518 long non-protein coding RNAs (lncRNAs). Gene Ontology (GO) analyses revealed that the top GO terms for *cis-*eGenes are enriched for immune functions (FDR <0.05). The *cis*-eQTL variants are enriched for SNPs reported to be associated with 815 traits in prior GWAS, including cardiovascular disease risk factors. As proof of concept, we used this eQTL resource in conjunction with genetic variants from public GWAS databases in causal inference testing (e.g., COVID-19 severity). After Bonferroni correction, Mendelian randomization analyses identified putative causal associations of 60 eGenes with systolic blood pressure, 13 genes with coronary artery disease, and seven genes with COVID-19 severity. This study created a comprehensive eQTL resource via BioData Catalyst that will be made available to the scientific community. This will advance understanding of the genetic architecture of gene expression underlying a wide range of diseases.

## INTRODUCTION

Over the past decade, genome-wide association studies (GWAS) have revolutionized understanding of the genetic architecture of complex traits.^1^ To date, GWAS have reported more than 59,000 associations (at *p*<5×10^−8^) between common genetic variants and numerous phenotypes (GWAS Catalog, v1.0.2).^2^ Yet, despite the clear success of GWAS, most single-nucleotide polymorphisms (SNPs) identified in GWAS reside in non-coding regions^3-5^ and do not illuminate causal mechanisms underlying SNP-trait associations.^5^ We posit that many of these trait-associated non-coding SNPs are likely to be involved in the regulation of gene expression.

Expression quantitative trait locus (eQTL) analysis seeks to identify genetic variants that affect the expression of local (*cis*) or distant (*trans*) genes (eGenes). Until recently, eQTL analysis has relied on high throughput microarray technologies and spawned a wave of genome-wide eQTL studies^6-11^ including a recent study from our group.^12^ These studies aided the understanding of the functional relevance of many GWAS results. Importantly, a hypothesis-free genome-wide eQTL approach permits the identification of new putatively functional loci without requiring previous knowledge of specific regulatory regions.

Most previous eQTL analyses were limited by small sample sizes and by the imprecision of microarrays. Newer technologies of RNA sequencing (RNA-seq) and whole genome sequencing (WGS) of DNA add greater precision and relevance to eQTL analyses. In conjunction with the National Heart, Lung, and Blood Institute’s (NHLBI) Trans-Omics for Precision Medicine (TOPMed) Program,^13^ the Framingham Heart Study (FHS) has obtained whole genome sequencing (WGS) in ∼6100 study participants to help understand the molecular basis of heart, lung, blood, and sleep disorders and to advance precision medicine. Among FHS participants with WGS, RNA-seq was obtained in 2622 participants. We conducted genome-wide eQTL analyses using high-precision genotypes obtained via WGS and gene expression levels from RNA-seq of whole blood. The primary objectives of this study were three-fold. Firstly, it sought to provide a scientific resource of *cis* and *trans* gene-level eQTL data to facilitate understanding of the genetic architecture of gene expression traits. Secondly, it was aimed to provide eQTL data for long noncoding RNAs (lncRNAs) that were not captured in prior array-based eQTL studies. Thirdly, it attempted to demonstrate the utility of the eQTL resource in causal inference analyses.

## RESULTS

Of the 2622 FHS participants in eQTL analyses, 720 participants were from the FHS Offspring cohort (mean age 71±8 years; 59% women) and 1902 were from the Third Generation cohort (mean age 47±8 years; 52% women) (**Table 1**). We used 19,624,299 SNPs with a minor allele count (MAC)□≥10 and 58,870 expression levels in association analyses to identify gene-level eQTLs.

**Table 1.**
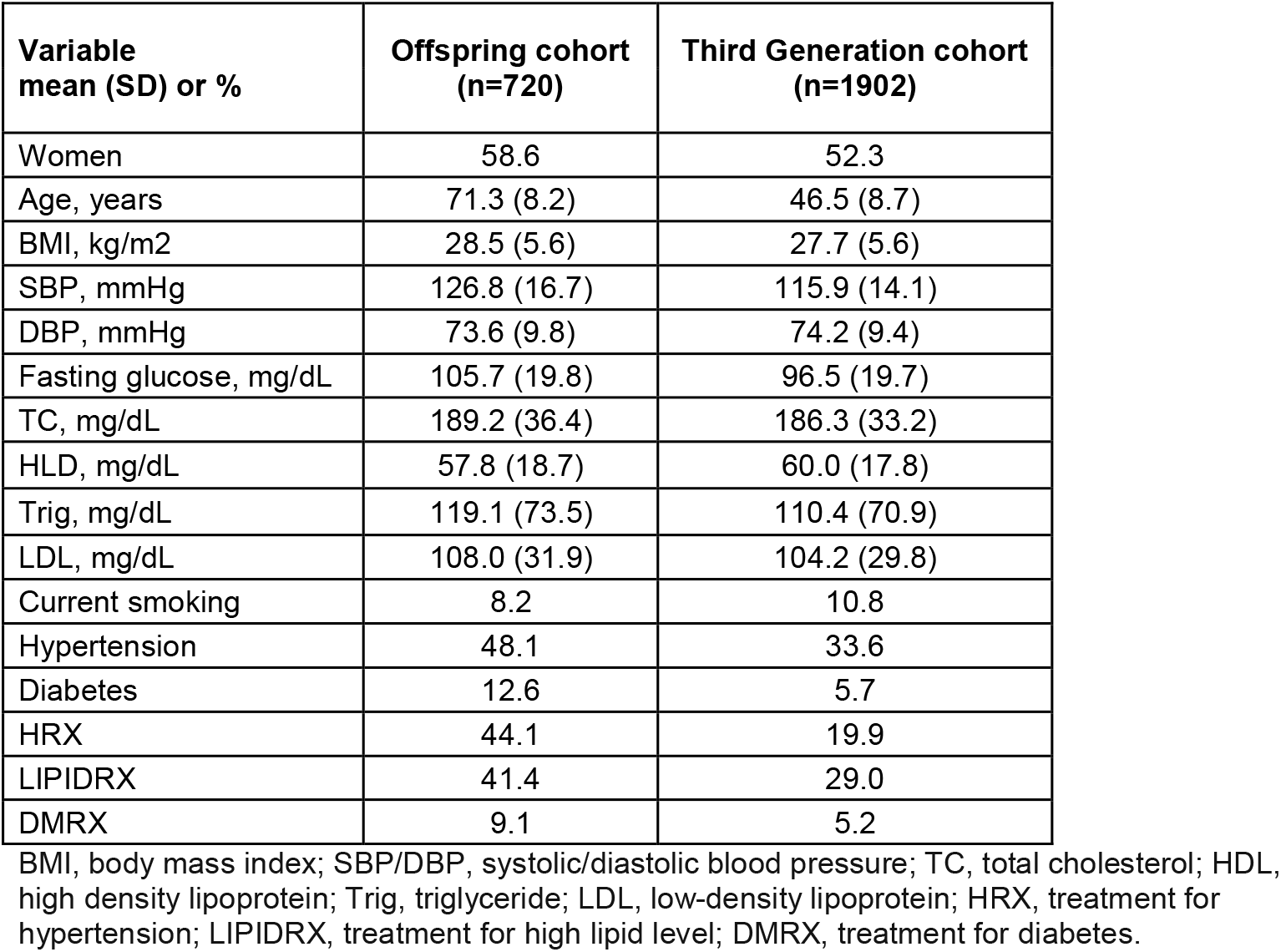
Participant characteristic

### Gene-level eQTL results

#### cis-eQTLs

*Cis*-eQTLs was defined as SNPs within 1 Mb of the transcription start sites (TSSs) of targeting genes. We identified 6,778,286 significant *cis*-eQTL variant-eGene pairs from 2,855,111 unique *cis*-eQTL variants and 15,982 unique eGenes (at *p*<5×10^−8^) (**Table 2**). The median number of *cis*-eQTL variants per gene was 183 (interquartile range=47,463). The eGenes harboring the largest numbers of *cis*-eQTL variants are located in the human leukocyte antigen (*HLA*) or major histocompatibility complex (MHC) on chromosome 6, reflecting a large number of SNPs in strong linkage disequilibrium (LD) at the MHC locus.^14^ Owing to the computational burden, we selected the strongest *cis*-eQTL variant (i.e., the lead variant) as that which had the lowest *p*-value per eGene. If several *cis*-eQTLs displayed the same *p*-value (i.e., they are in perfect LD, r^2^=1), we randomly select one lead eQTL variant per eGene (**Table 3** & **Supplemental Table 1**). Of the 6,778,286 significant *cis*-eQTL variant-eGene pairs, 82.8% (n=13,226) of SNPs were within 100 kb of the TSSs of the respective eGenes, 9.3% (n=1492) within 101 kb – 200 kb region, 5.7% (n=910) within 201 kb – 500 kb region, and 2.2% (n=352) within 501 kb – 1 Mb (**Figure 1**). Among the selected lead *cis*-eQTL variants, 85% (n=13584) explained a small proportion of variation (R^2^ < 0.2) in expression of the respective eGenes, 197 (1.2%) and 27 (0.17%) of lead *cis*-eQTL variants explained a moderately large (R^2^ 0.6 to 0.8) or a very large proportion of variation in expression (R^2^>0.80) of the corresponding eGenes (**Figure 1**).

**Table 2.** *Cis*- and *trans*-eQTL in the Framingham Heart Study

|  | eQTLs at gene level | lncRNA eQTLs |
| --- | --- | --- |
| Cis-eQTL-eGene ($p < 5e-8$) | | |
| Number of pairs | 6,778,286 pairs<br>2,855,111 unique cis-eQTLs and 15,982 eGenes | 442,379 *cis*-eQTLs are located in 1518 *cis*-lncRNAs genes |
| Trans-eQTL-eGene ($p < 1e-12$) | | |
| Number of pairs | 1,469,754 pairs<br>526,056 unique trans-eQTLs and 7,233 trans-eGenes | 117,862 *trans*-eQTLs are located in 475 *trans*-lncRNAs genes |

**Table 3.** Top 25 cis-eQTLs (*p*<5e-8)

| Gene Symbol | SNP | Chr | SNP position | Gene start position | R<sup>2</sup> | Beta | log10P | OA | EA | EAF | Type |
| --- | --- | --- | --- | --- | --- | --- | --- | --- | --- | --- | --- |
| PPIE | rs7513045 | 1 | 39738494 | 39692182 | 0.84 | 11.40 | -1029.25 | G | T | 0.36 | protein_coding |
| CCDC163 | rs4660860 | 1 | 45480561 | 45493866 | 0.90 | -3.10 | -1286.89 | T | A | 0.30 | protein_coding |
| CYP26B1 | rs13430651 | 2 | 72215195 | 72129238 | 0.81 | 1.98 | -920.005 | G | A | 0.15 | protein_coding |
| MAP3K2-<br>~T | rs2276683 | 2 | 127389186 | 127389130 | 0.88 | -1.61 | -1176.37 | G | C | 0.23 | lincRNA |
| SLC12A7 | rs35188965 | 5 | 1104823 | 1050384 | 0.81 | -29.87 | -915.459 | C | T | 0.44 | protein_coding |
| ENC1 | rs112772452 | 5 | 74631048 | 74627406 | 0.83 | 14.53 | -986.798 | CA | C | 0.11 | protein_coding |
| ERAP2 | rs2910686 | 5 | 96916885 | 96875939 | 0.85 | 36.98 | -1044.91 | T | C | 0.43 | protein_coding |
| BTNL3 | rs72494581 | 5 | 181003797 | 180988845 | 0.82 | 13.52 | -950.405 | T | C | 0.30 | protein_coding |
| HLA-DRB5 | rs68176300 | 6 | 32558713 | 32517353 | 0.83 | -178.13 | -1003.76 | T | G | 0.15 | protein_coding |
| AL512625.3 | rs1845054 | 9 | 62906092 | 62856999 | 0.83 | -1.19 | -993.655 | T | C | 0.13 | lincRNA |
| CUTALP | rs13299616 | 9 | 120832525 | 120824828 | 0.86 | -23.25 | -1092.88 | T | C | 0.40 | transcribed_unitary_pseudogene |
| LDHC | rs201993031 | 11 | 18412985 | 18412318 | 0.82 | 0.16 | -946.833 | CCCTTCCTT | C | 0.12 | protein_coding |
| ACCS | rs2074038 | 11 | 44066439 | 44065925 | 0.83 | 16.69 | -997.26 | G | T | 0.11 | protein_coding |
| FADS2 | rs968567 | 11 | 61828092 | 61792980 | 0.88 | 31.41 | -1186.37 | C | T | 0.17 | protein_coding |
| XRRA1 | rs10899051 | 11 | 74931506 | 74807739 | 0.91 | 5.38 | -1327.88 | G | A | 0.26 | protein_coding |
| B4GALNT3 | rs1056008 | 12 | 553672 | 460364 | 0.85 | 6.71 | -1043.34 | T | C | 0.25 | protein_coding |
| DDX11 | rs3891006 | 12 | 31073506 | 31073860 | 0.86 | -13.25 | -1102.08 | A | G | 0.44 | protein_coding |
| RPS26 | rs1131017 | 12 | 56042145 | 56041351 | 0.81 | -134.34 | -929.902 | C | G | 0.39 | protein_coding |
| C17orf97 | rs7503725 | 17 | 410351 | 410325 | 0.85 | 1.89 | -1055.68 | G | T | 0.25 | protein_coding |
| AC126544.2 | rs2696531 | 17 | 46278268 | 45586452 | 0.86 | 1.04 | -1097.79 | C | A | 0.21 | lincRNA |
| SPATA20 | rs9890200 | 17 | 50547162 | 50543058 | 0.81 | -11.01 | -934.173 | A | C | 0.37 | protein_coding |
| CEACAMP3 | rs3745936 | 19 | 41586462 | 41599735 | 0.84 | 1.11 | -1040.05 | A | T | 0.22 | transcribed_unprocessed_pseudogene |
| PWP2 | rs2277806 | 21 | 44089769 | 44107373 | 0.87 | 3.16 | -1139.85 | A | C | 0.19 | protein_coding |
| GATD3A | rs3788104 | 21 | 44092213 | 44133610 | 0.86 | 4.25 | -1104.35 | G | A | 0.18 | protein_coding |
| FAM118A | rs576259663 | 22 | 45363712 | 45308968 | 0.86 | 43.45 | -1108.47 | T | TA | 0.12 | protein_coding |

EA, effect allele; OA, the other allele

**Figure 1.**
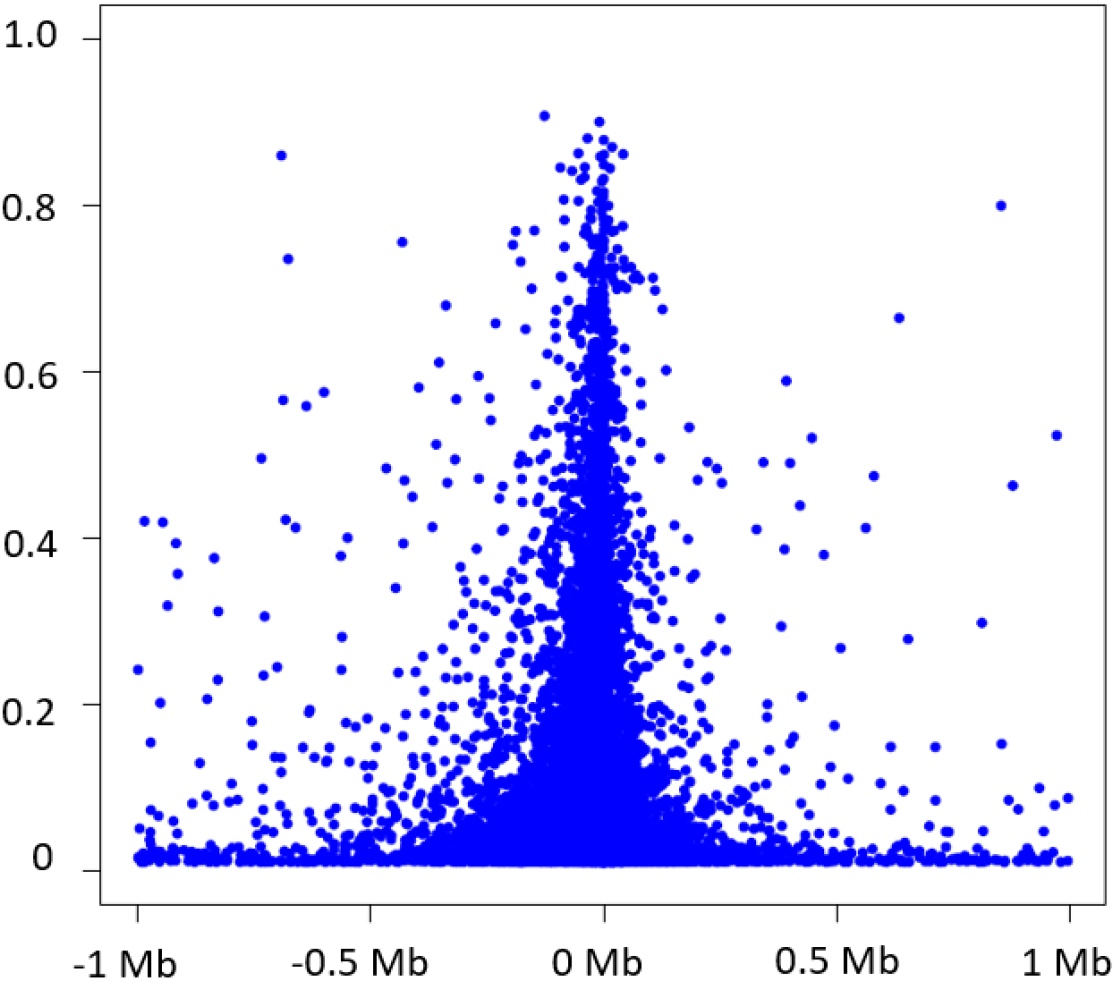
Variance in eGenes explained by significant cis-eQTLs in relation to the distance of significant cis-eQTLs to the transcription start site of the cis-gene.

#### trans-eQTLs

*Trans*-eQTLs referred to the SNPs that were beyond of 1 Mb of the TSSs of the eGenes on the same chromosome or those on the different chromosomes of the eGenes. We identified 1,469,754 significant *trans*-eQTL variant-eGene pairs (*p*<1e-12) from 526,056 unique *trans*-eQTL variants and 7,233 *trans*-eGenes (**Table 2**). The median number of significant-eQTL variants per eGene was 11 (interquartile range=2, 76).^14^ With the same method used to select the lead *cis*-eQTL variants, we selected the lead trans-eQTL variant based on *p*-values for each *trans*-eGene (**Supplemental Table 2**). The top 25 *trans*-eQTL are listed in **Table 4**. Among the lead *trans*-eQTL variants, 95.8% (n=6926) explained a small proportion of variation in expression (R^2^ < 0.2) of the corresponding eGenes, 27 (0.37%) and five (0.07%) lead *trans*-eQTL variants explained a moderately large (R^2^ in 0.6 to 0.8) or a very large (R^2^ >0.80) proportion of variation in expression of the corresponding *trans*-eGenes. The *trans*-eQTL variants, rs1442867716 (*GATD3A*), rs74987185 (*RPSAP58*), rs538628 (*AC126544*.*2*), rs16997659 (*EIF2S3B*), rs3927943 (*NPIPB15*) explained >0.8 of variance in the expression of their respective *trans-*eGenes.

**Table 4.** Top 25 top trans-eQTLs (*p*< 1e-12)

| Gene Symbol | SNP | Gene Chr | SNP Chr | SNP Pos | Gene Start Pos | R<sup>2</sup> | Beta | t value | log10P | OA | EA | EAF | Gene type |
| --- | --- | --- | --- | --- | --- | --- | --- | --- | --- | --- | --- | --- | --- |
| EMBP1 | rs4549528 | 1 | 5 | 50372700 | 121519112 | 0.70 | 1.63 | 78.04 | -677.53 | T | C | 0.48 | transcribed_unprocessed_pseudogene |
| AL365357.1 | rs4841 | 1 | 5 | 150446963 | 178411616 | 0.64 | 2.79 | 67.64 | -570.81 | C | T | 0.25 | processed_pseudogene |
| AL591846.1 | rs13161099 | 1 | 5 | 150442799 | 206695837 | 0.62 | 1.87 | 65.67 | -549.988 | G | A | 0.25 | processed_pseudogene |
| AC004057.1 | rs1131017 | 4 | 12 | 56042145 | 113214046 | 0.61 | -0.42 | -63.03 | -521.823 | C | G | 0.39 | transcribed_processed_pseudogene |
| RPL10P9 | rs6655287 | 5 | X | 154396528 | 168616352 | 0.64 | 4.11 | 68.52 | -580.03 | A | G | 0.10 | processed_pseudogene |
| PSPHP1 | rs34945686 | 7 | 7 | 65809663 | 55764797 | 0.61 | 0.05 | 64.11 | -533.329 | C | G | 0.18 | unprocessed_pseudogene |
| AC104692.2 | rs6593279 | 7 | 7 | 55736277 | 152366763 | 0.60 | 0.05 | 62.97 | -521.195 | G | A | 0.20 | processed_pseudogene |
| RNF5P1 | rs8365 | 8 | 6 | 32180626 | 38600661 | 0.78 | 0.97 | 96.24 | -850.788 | G | C | 0.19 | processed_pseudogene |
| TUBB8 | rs28652789 | 10 | 16 | 33807 | 46892 | 0.61 | 0.32 | 63.35 | -525.289 | G | C | 0.25 | protein_coding |
| COX20P1 | rs10927332 | 10 | 1 | 244837362 | 68632371 | 0.62 | 0.10 | 64.57 | -538.221 | C | T | 0.19 | processed_pseudogene |
| EIF2S3B | rs16997659 | 12 | X | 24057745 | 10505602 | 0.81 | 0.99 | 106.39 | -939.701 | A | G | 0.17 | protein_coding |
| RPS2P5 | rs2286466 | 12 | 16 | 1964282 | 118246084 | 0.80 | 71.71 | 101.17 | -894.683 | A | G | 0.21 | processed_pseudogene |
| LINC00431 | rs41288614 | 13 | 13 | 112486035 | 110965704 | 0.70 | 0.20 | 76.92 | -666.36 | A | G | 0.15 | transcribed_unprocessed_pseudogene |
| NPIPB15 | rs3927943 | 16 | 16 | 69977282 | 74377878 | 0.80 | 3.79 | 103.12 | -911.688 | T | A | 0.40 | protein_coding |
| TUBB8P7 | rs28652789 | 16 | 16 | 33807 | 90093154 | 0.75 | 0.51 | 88.54 | -779.687 | G | C | 0.25 | transcribed_unprocessed_pseudogene |
| RPL13P12 | rs2280370 | 17 | 16 | 89561052 | 17383377 | 0.69 | 36.16 | 75.78 | -654.808 | T | G | 0.19 | processed_pseudogene |
| LRRC37A2 | rs56328224 | 17 | 17 | 45495053 | 46511511 | 0.80 | 5.91 | 101.76 | -899.821 | C | T | 0.24 | protein_coding |
| POLRMTP1 | rs14155 | 17 | 19 | 619021 | 62136972 | 0.69 | 0.62 | 75.32 | -650.176 | G | C | 0.50 | processed_pseudogene |
| TUBB8P12 | rs2562131 | 18 | 16 | 33887 | 47390 | 0.65 | 0.47 | 68.64 | -581.244 | C | A | 0.25 | protein_coding |
| AP001005.3 | rs28652789 | 18 | 16 | 33807 | 49815 | 0.61 | 0.15 | 64.25 | -534.859 | G | C | 0.25 | lincRNA |
| RPSAP58 | rs74987185 | 19 | 3 | 39414963 | 23827162 | 0.84 | 10.17 | 117.60 | -1031.88 | G | GCT | 0.31 | processed_pseudogene |
| GATD3B | rs2277806 | 21 | 21 | 44089769 | 5079294 | 0.74 | -3.83 | -84.78 | -743.85 | A | C | 0.19 | protein_coding |
| FP565260.1 | rs2277806 | 21 | 21 | 44089769 | 5130871 | 0.76 | -2.96 | -90.65 | -799.469 | A | C | 0.19 | protein_coding |
| SIRPAP1 | rs115287948 | 22 | 20 | 1915413 | 30542536 | 0.75 | 1.12 | 89.28 | -786.711 | G | A | 0.36 | processed_pseudogene |
| GPX1P1 | rs7643586 | X | 3 | 49394214 | 13378735 | 0.61 | 16.44 | 64.25 | -534.823 | C | G | 0.43 | processed_pseudogene |

EA, effect allele; OA, the other allele

#### Long noncoding RNA (lncRNA) eQTLs

lncRNAs are usually more than 200 bases in length, share no conserved sequence homology, and have variable functions.^15^ Of the 58,870 transcripts captured by RNA-seq 7696 (13%) are lncRNAs. Of the significant *cis*-eQTL variant-eGene pairs (*p*<5e-8), 447,598 *cis*-eQTL variants are associated with expression of 1518 unique *cis*-lncRNAs. The top *cis-*eQTL-lncRNA variant-gene pairs are listed in **Supplemental Table 3**. Of the significant *trans*-eQTL variant-eGene pairs (*p* < 1e-12), 121,241 *trans*-eQTL variants were associated with expression of 475 *trans*-lncRNAs. The top *trans-*eQTL-lncRNA variant-gene pairs are listed in **Supplemental Table 4**. Three *cis*-eQTL-lncRNA pairs were observed among the top 25 *cis*-eQTL results (**Table 3**). The top *cis*-lncRNA, the MAP3K2 divergent transcript (MAP3K2-DT), is the only lncRNA that is located adjacent to a protein coding gene, the 5’-end of mitogen-activated protein kinase kinase kinase 2 (MAP3K2) on chromosome 2 (q14.3) (**Figure 2**). The correlation of expression of expression of MAP3K2 and MAP3K2-DT was weak (Pearson correlation = 0.08; *p =* 0.12). Among the top 25 *trans*-eQTL pairs, we identified one *trans*-eQTL-lncRNA pair (**Table 4**). The top *trans-*lncRNA, AP001005.3 on chromosome 18, is not adjacent to any known genes.

**Figure 2.**
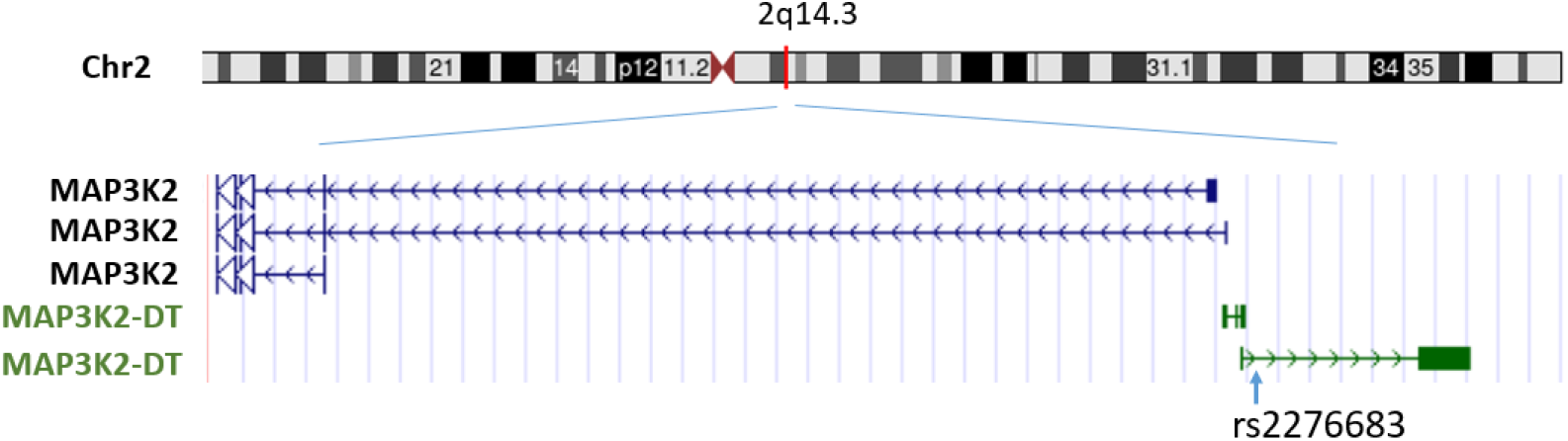
Cis-long noncoding RNA, MAP3K2-DT, and the lead cis-eQTL, rs2276683.

### Gene Ontology analyses

We identified 100 significant GO terms for the top 1000 *cis*-eGenes at FDR < 0.05. Of these Go terms, there were 58 for Biological Process, 31 for Cellular Component, and 11 for Molecular Function (**Supplemental Table 5**). Of note, the top GO terms appeared to be related to immune functions. For example, the top two Biological Processes are “leukocyte degranulation” (FDR = 1e-6) and “myeloid leukocyte mediated immunity” (FDR = 2e-6) and the top two Cellular Components are cytoplasm (FDR = 3e-6) and MHC protein complex (FDR = 6e-6). The top 1000 top *trans*-eGenes gave rise to 75 significant (FDR < 0.05) GO terms including 37 for Biological Process, 32 for Cellular Component, and 6 for Molecular Function. The top GO terms for the top 1000 *trans*-eGenes were enriched in pathways and molecular functions related to immune functions (**Supplemental Table 5**).

### GWAS enrichment analyses

We linked 1,855,111 *cis*-eQTL variants (*P* <5e-8) to GWAS Catalog variants. At FDR < 0.05, the *cis*-eQTL variants were enriched with GWAS SNPs associated with 815 traits, representing 28% of the traits in the GWAS Catalog. The top traits identified in enrichment analyses include several cardiovascular disease risk factors. For example, *cis*-eQTL variants are enriched with BMI-associated SNPs (fold enrichment=84, FDR = 3.3e-267), total cholesterol (fold enrichment=98, FDR=7.3e-162) (**Supplemental Table 6**). We identified 193 GWAS traits enriched for the *trans*-eQTL variants (**Supplemental Table 7**). The top traits in the *trans* enrichment analysis included neuroticism measurement (fold enrichment=3, FDR=1.9e-89) and BMI-adjusted waist circumference (fold enrichment=2, 6.4e-87).

### Mendelian randomization analysis

We performed two-sample MR to test for potential causal association of the *cis*-eGenes with SBP, CAD, and COVID-19 severity. We found 1558 genes containing at least one eQTL variant (median 29; interquartile range [IQR] 6, 88) that coincided with variants from GWAS of SBP (*p* < 5e-8).^16^ After Bonferroni correction for multiple testing, MR identified putative causal associations for 60 genes with SBP (i.e., *p*<0.05/1558) (**Table 5** & **Supplemental Table 8**). Of these 60 genes, six lncRNAs (AC066612.1, AC069200.1, AC092747.4, AC100810.3, AL590226.2, and LY6E-DT) showed putative causal associations with SBP. For CAD, 173 genes contained at least one eQTL variant [median 5; IQR (2, 18) that also were associated with CAD in GWAS.^17^ Thirteen genes showed putative causal associations with CAD (i.e., *p* < 0.05/173) (**Table 5** & **Supplemental Table 8**); none of the 13 putative causal genes was a lncRNA. Using results of a recent GWAS of COVID-19 severity^18^ and a study that investigated circulating proteins influencing COVID-19 susceptibility and severity,^19^ we identified 24 genes with *cis*-eQTL variants [median 3, IQR; (2, 126)] that coincide with COVID severity variants. MR analyses identified seven putatively causal genes for COVID-19 severity (**Table 5** and **Supplemental Tables 8 & 9**). Two of the genes included the 2’-5’-oligoadenylate synthetase 1 gene (*OAS1*) (MR IVW *p* =1.6E-04) and the interferon-alpha/beta receptor beta chain gene (*IFNAR2*) (MR IVW *p* = 1.8E-06). A recent study identified an alternative splicing variant (sQTL), rs10774671, at exon 7 of *OAS1* for which the “G” allele leads to a “prenylated” protein that is protective against severe COVID.^20^ Additional MR analysis using rs10774671 as the instrumental variable demonstrated that splice variation of *OAS1* is also causal for COVID-19 severity (*p* =4e-6).

**Table 5.** Top results in Mendelian randomization analyses

| Exposure | Chr | Gene type | Outcome | INV MR<sup>1</sup> |  |  | N SNPs |
| --- | --- | --- | --- | --- | --- | --- | --- |
|  |  |  |  | Beta | SE | p |  |
| *PSRC1* | 1 | Protein coding | CHD | -0.084 | 0.0075 | 4.8E-29 | 7 |
| *LTA* | 6 | Protein coding | CHD | -0.069 | 0.011 | 1.3E-09 | 5 |
| *MIR6891* | 6 | miRNA | CHD | 1.72 | 0.28 | 2.0E-09 | 25 |
| *LIPA* | 10 | Protein coding | CHD | 0.0033 | 0.00039 | 2.9E-17 | 18 |
| *PHETA1* | 12 | Protein coding | CHD | -0.078 | 0.013 | 4.7E-09 | 3 |
| *ACSL6* | 5 | Protein coding | COVID-19 | 0.19 | 0.064 | 0.0025<sup>#</sup> | 4 |
| *DPP9* | 19 | Protein coding | COVID-19 | -0.044 | 0.017 | 0.0078<sup>#</sup> | 3 |
| *HLA-DRB1* | 6 | Protein coding | COVID-19 | 0.00099 | 0.00018 | 1.9E-08<sup>#</sup> | 35 |
| *IFNAR2* | 21 | Protein coding | COVID-19 | -0.023 | 0.0037 | 1.8E-06<sup>#</sup> | 11 |
| *OAS1* | 12 | Protein coding | COVID-19 | -0.0086 | 0.0022 | 1.6E-04<sup>\$</sup> | 1 |
| *SLC22A31* | 12 | Protein coding | COVID-19 | 0.32 | 0.11 | 0.0029 | 13 |
| *TYK2* | 21 | Protein coding | COVID-19 | 0.011 | 0.0021 | 2.8E-08 | 3 |
| *AC006460.2* | 2 | Bidirectional promoter lncRNA | SBP | -5.60 | 0.55 | 2.3E-24 | 3 |
| *MAP4* | 3 | Protein coding | SBP | 0.092 | 0.0086 | 4.6E-27 | 4 |
| *PHETA1* | 12 | Protein coding | SBP | -0.92 | 0.058 | 1.9E-58 | 3 |
| *SLC5A11* | 16 | Protein coding | SBP | -0.82 | 0.066 | 5.3E-35 | 21 |
| *ACADVL* | 17 | Protein coding | SBP | -0.035 | 0.0030 | 1.5E-31 | 3 |

<sup>1</sup>, Beta/SE and p-value were obtained by inverse variance weighted MR method.
<sup>#</sup>, Heterogeneity was observed in MR analyses. Sensitivity analyses were performed with median-based and mode-based MR methods in Supplemental Table 9.
<sup>\$</sup>, MR analysis was performed at gene level. At splice variation level (rs10774671), the MR $p = 4\text{E-}06$ .

### Replication analyses

Of the reported 10,914 *cis*-eQTL-eGene pairs from the study by Battle et al. ^21^ (FDR < 0.05), ^21^ 6782 (62%) pairs displayed *p* < 5e-8 in the present study. The average proportion of variance explained by these 6782 *cis*-eQTL variants in respective genes was 0.11 (**Supplemental Table 10)**. Of the 269 *trans*-eQTL-eGene pairs (FDR < 0.05) reported by Battle et al. ^21^ 47 (18%) pairs displayed *p* < 1e-12 in the current study. The average proportion of variance explained by these 47 *trans*-eQTL variants in respective genes was 0.076. Of note, all 47 *trans*-eQTL variants and respective *trans*-eGenes are located on the same chromosomes (**Supplemental Table 11**). The average distance between these *trans*-eQTL variants and respective *trans*-eGenes is within 22 Mb.

We conducted additional replication analysis for the *cis*-eQTL variant-eGene pairs generated from 8,372,247 SNPs and 20,188 gene transcripts that were common to our study (n = 2622 participants) and to GTEx(6) (n = 755 participants) (**Supplemental Figure 1**). At *p* < 5e-8, we identified 1,080,485 *cis*-eQTL variant-eGene pairs in GTEx and 3,852,182 pairs in our study; of these, 951,085 pairs (88% of pairs in GTEx) displayed the same effect direction as in our larger study. At *p* < 1e-4, we identified 1,815,208 *cis*-eQTL variant-eGene pairs in GTEx and 6,364,173 pairs in this study; of these, 1,797,977 (99% of pairs in GTEx) displayed the same effect directionality with our study (**Supplemental Figure 1**).

## DISCUSSION

We leveraged WGS and RNA-seq data from 2,622 FHS participants to create a powerful scientific resource of eQTLs. We identified significant unique *cis*-eQTL variants-eGene pairs (*n* = 2,855,111 unique variants with *cis*-15,982 eGenes) and 526,056 unique *trans*-eQTL variants-eGene pairs (526,056 unique variants and unique 7,233 *trans*-eGenes. A large proportion of reported *cis*-eQTL variant-eGene pairs were replicated with directionally concordant in our study including 88% of *cis*-variant-eGene pairs from GTEx.

Consistent with our previous study and others, ^7-12,22,23^ 90% of eQTL variants identified in the present study are located in within 1 Mb of the corresponding cis-eGene and 83% are within 100 kb of the TSSs of the corresponding eGene. While the majority of (85% of *cis-* and 96% of *trans-*) lead eQTL variants explained only a small proportion (R^2^ <0.2) of interindividual variation in expression of the corresponding eGenes, 15% of lead *cis*-eQTL variants and 4% of lead *trans* variant explained 20% or more of interindividual variation in expression of the corresponding eGenes ^24^. Additionally, eQTL variants were enriched (*p*<0.0001) in disease-associated SNPs identified by GWAS. We further demonstrated the utility of our eQTL resource for conducting causal inference testing. Our MR analyses revealed putatively causal relations of gene expression to several disease phenotypes including SBP, CAD, and COVID-19 severity. Taken together, the comprehensive eQTL resource we provide can advance understanding of the genetic architecture of gene expression underlying a wide variety of diseases. The interactive and browsable eQTL resource will be posted to the National Heart, Lung, and Blood Institute’s BioData Catalyst site and will be freely accessible to the scientific community.

Our study expands current knowledge by creating an accessible and browsable resource of eQTLs based on WGS and RNA-seq technologies. It also includes eQTLs for lncRNAs that were not reported in prior eQTL studies that used array-based expression profiling. Over the past decade, accumulating evidence shows that lncRNAs are widely expressed and have key roles in gene regulation.^25,26^ It is estimated that the human genome contains 16,000 to 100,000 lncRNAs.^25^ We identified 447,598 *cis*-eQTL variants for 1518 *cis*-lncRNAs and 121,241 *trans*-eQTLs for 475 *trans*-lncRNAs (**Supplemental Tables 3&4**). In addition, we identified six lncRNAs that showed putative causal associations with SBP. However, the functions of these six lncRNAs remain to be determined. Thus, our novel eQTL database may also help in the study of non-protein-coding RNAs in relation to health and disease.

As a proof of concept of the application of the eQTL resource, we performed MR analyses on a small number of cardiovascular traits and COVID-19 severity and demonstrated that the eQTL database can identify promising candidate genes with evidence of putatively causal relations to disease that may merit functional studies. Severe acute respiratory syndrome coronavirus 2 (SARS-CoV-2) has spread across the globe and caused millions of deaths since it emerged in 2019. Recent GWAS of COVID-19 susceptibility and severity ^27-29^ have identified SNPs in several loci on chromosomes 3, 9 and 21.^30^ Using our eQTL resource in conjunction with COVID-19 GWAS, we conducted MR analyses that identified seven genes, including *OAS1* and *IFNAR2*, as putatively causal for COVID-19 severity. The *OAS1/2/3* cluster has been identified as a risk locus for COVID-19 severity.^27^. This area harbors a protective haplotype of approximately 75 kilo-bases (kb) at 12q24.13 among individuals of European ancestry.^19^ A recent study identified an alternative splicing variant, rs10774671, at exon 7 of *OAS1* for which the protective allele “G” leads to a more active OAS1 enzyme.^20^ Our MR results suggest that both the *OAS1* gene expression level and its splice variation are causal for COVID-19 severity.

The *IFNAR2* gene encodes a protein in the type II cytokine receptor family. Mutations in *IFNAR2* are associated with Immunodeficiency and measles virus susceptibility and play an essential and a narrow role in human antiviral immunity.^31^ A recent study further showed that loss-of-function mutations in *IFNAR2* are associated with severe COVID-19.^32^ These studies, considered alongside our MR results provide evidence of a causal role of *IFNAR2* expression in severe COVID-19 infection.

This study has several noteworthy limitations. This study included White participants of European ancestry who were middle-aged and older; therefore, the eQTLs identified may not be generalizable to other races or age ranges. The current RNA-seq platform included ∼7700 lncRNAs, which is a modest subset of all lncRNAs in the human genome.^25^ We used MR analyses to infer causal relation of genes to disease traits. MR analysis is predicated on a set of critical assumptions that may not be testable in the setting of eQTL analysis.^33,34^ Replication of our eQTL findings is warranted in studies with larger sample sizes and more diverse populations.

Our study also has several strengths. The advent of high-throughput RNA sequencing technology provides an unparalleled opportunity to accelerate understanding of the genetic architecture of gene expression. Our study extends and expands the existing literature by identifying novel eQTLs based on WGS and RNA-seq. We demonstrate the potential applications of a vast eQTL resource by analyzing the concordance of eQTL variants with SNPs from GWAS of several disease phenotypes followed by causal inference analyses that identified promising disease-related genes that may merit functional studies. We created an open and freely accessible eQTL repository that can serve as a promising scientific resource to better understand of the genetic architecture of gene expression and its relations to a wide variety of diseases.

## METHODS

### Study participants

This study included participants from the FHS Offspring [10] and Third Generation cohorts [11]. Blood samples for RNA seq were collected from Offspring participants who attended the ninth examination cycle (2011–2014) and the Third Generation participants who attended the second examination cycle (2008–2011). Protocols for participant examinations and collection of genetic materials were approved by the Institutional Review Board at Boston Medical Center. All participants provided written, informed consent for genetic studies. All research was performed in accordance with relevant guidelines/regulations.

### Isolation of RNA from whole blood and RNA-seq

Peripheral whole blood samples (2.5 mL) were collected from FHS participants (Offspring participants at the ninth examination cycle and the Third Generation participants at the second examination cycle) using PAXgene(tm) tubes (PreAnalytiX, Hombrechtikon, Switzerland), incubated at room temperature for 4 hours for RNA stabilization, and then stored at -80 °C until use. Total RNA was isolated using a standard protocol using a PAXgene Blood RNA Kit at the FHS Genetics Laboratory (FHS Third Generation cohort) and the TOPMed contract laboratory at Northwest Genomics Center (Offspring cohort). Tubes were allowed to thaw for 16 hours at room temperature. White blood cell pellets were collected after centrifugation and washing. Cell pellets were lysed in guanidinium-containing buffer. The extracted RNA was tested for its quality by determining absorbance readings at 260 and 280 nm using a NanoDrop ND-1000 UV spectrophotometer. The Agilent Bioanalyzer 2100 microfluidic electrophoresis (Nano Assay and the Caliper LabChip system) was used to determine the integrity of total RNA.

All RNA samples were sequenced by an NHLBI TOPMed program ^13^ reference laboratory (Northwest Genomics Center) following the TOPMed RNA-seq protocol. All RNS-seq data were processed by University of Washington. The raw reads (in FASTQ files) were aligned using the GRCh38 reference build to generate BAM files. RNA-SeQC^35^ was used for processing of RNA-seq data by the TOPMed RNA-seq pipeline to derive standard quality control metrics from aligned reads. Gene-level expression quantification was provided as read counts and transcripts per million (TPM). GENCODE 30 annotation was used for annotating gene-level expression.

### Whole blood cell counts

Whole blood cell counts include white blood cell (WBC) count, red blood cell count, platelet count, and WBC differential percentages (neutrophil percent, lymphocyte percent, monocyte percent, eosinophil percent, and basophil percent). Contemporaneously measured blood cell counts were available in 2094 (80%) of the 2622 FHS participants used in eQTL analyses. We performed partial least squares (PLS) prediction method^36^ with three-fold cross-validation (2/3 samples for training and 1/3 for validation) to impute these blood cell components using gene expression from RNA-seq. Prediction accuracy (R-squared) varied across blood component: WBC: 0.58, platelet: 27%, neutrophil percentage: 82%, lymphocyte percentage: 85%, monocyte percentage: 77%, eosinophil percentage: 87%, basophil percentage: 32%. Because 80% of the participants in this study had directly measured cell count variables and only 20% received imputed variables, we used the measured (in 2094 participants) and predicted (in 528 participants) blood cell components as covariates in regression models for eQTL analyses.

### RNA-seq quality control, and data adjustment

To minimize confounding, expression residuals were generated by regressing transcript expression level on age, sex, measured or predicted blood cell count and differential cell proportions, and genetic principal components. Principal component (PC) analysis is a technique for reducing the dimensionality in large data sets. ^37^ It has been widely used in regression analyses to minimize unknown confounding. We included five PCs computed from FHS genotype profiles to account for population stratification. We also included 15 PCs computed from the transcriptome profile to account for unknown confounders that may affect gene expression. In addition, we adjusted for a relatedness matrix, and technical covariates including year of blood collection, batch (sequencing machine and time, plate and well), and RNA concentration.

### Whole genome sequencing

Whole genome sequencing of genomic DNA from whole blood was conducted in ∼6,000 FHS participants as part of NHLBI’s TOPMed program.^13^ Standard procedures were used to obtain DNA fragmentation and library construction. Sequencing was performed by a TOPMed reference laboratory (the Broad Institute of MIT and Harvard) using HiSeq X with sequencing software HiSeq Control Software (HCS) version 3.3.76, then analyzed using RTA2 (Real Time Analysis). The DNA sequence reads were aligned to a human genome build GRCH38 using a common pipeline across all TOPMed WGS centers. A sample’s sequence was considered complete when the mean coverage of nDNA was ≥30x. This analysis used genetic variants generated from TOPMed Freeze 10a.^13^

### Association analyses of expression levels with SNPs

We performed association analyses of expression levels with genome-wide SNPs with minor allele frequencies (MAFs) ≥ 0.01. In a simple regression model, a SNP was used as an independent variable and the residuals of a transcript expression level was used as the dependent variable. All analyses were performed on the NIH-supported STRIDES cloud infrastructure. A graphical Processing Unit (GPU)-based program ^12^ was used to facilitate computation. Effect sizes, standard error, partial R-squared, and p-values for all SNP-gene expression pairs were stored to enable complete lookups and to facilitate later meta-analysis. In this study, we defined *cis*-eQTLs as targeting genes within 1 Mb of their transcription start site (TSS). *Trans*-eQTLs referred to those that were beyond of 1 Mb of the TSSs of the eGenes on the same chromosome or those on the different chromosomes of the eGenes. A significant *cis*-eQTL of an eGene was identified if a SNP within 1 Mb of that gene was associated with expression of a transcript of that gene at *P* < 5×10^−8^. A significant *trans-*eQTL was defined as a SNP beyond 1 Mb that gave rise to *P* < 1×10^−12^ in association a gene.

### Gene Ontology analyses

We selected the single, most significant eQTL variant (i.e. lead variant) for each eGene (for the gene level analysis) from *cis*- and *trans*-eQTL results separately. The eGenes annotated to the selected lead *cis* and *trans* eQTL variants were matched into Entrez IDs. We used the “goana” function from the “limma” package^38^ to test for over-representation of gene ontology (GO) terms or KEGG pathways applied to the top 1000 eGenes. We used FDR < 0.05 to report GO terms including Biological Process, Cellular Component, and Molecular Function.

### Enrichment analyses using GWAS Catalog

We linked the eQTL variants with SNPs from the GWAS Catalog ^2^ (data downloaded on October 22, 2021), which included 243,618 entries for 2,960 mapped traits at p<5e-8. *Cis*- and *trans*-eQTL variants were analyzed separately. Unique SNP RS IDs were used for enrichment analysis with Fisher’s test. FDR < 0.05 was used for significance.

### Correlation analysis of selected lncRNA and protein coding genes

For lncRNAs that were in the top 25 *cis*-eQTL variant-eGene pairs, we performed partial Pearson correlation analyses between the expression level of the lncRNA and its nearby protein coding gene, adjusting for the same set of covariates that were included in eQTL analysis. We performed random sampling of 1000 genes 500 times to derive null distributions of partial Pearson correlation of these gene pairs. We calculated an empirical p-value to evaluate whether the partial Pearson correlation coefficient between the expression level of an lncRNA and its nearby protein coding gene was significantly higher than the average partial Pearson correlation coefficient from randomly selected gene pairs. The empirical p-value was calculated as the proportion of partial Pearson correlation coefficients that were more extreme than the correlation coefficient of an lncRNA and its nearby protein coding gene.

### Mendelian randomization analysis

We conducted Mendelian randomization (MR) to demonstrate the application of the eQTL resource in causal inference analysis. We tested for potential causal association of the *cis*-eGenes with SBP, coronary artery disease (CAD), and COVID-19 severity. SBP-associated SNPs were obtained from GWAS of over 1 million people.^16^ CAD-associated SNPs were obtained from the study of 34,541 CAD cases and 261,984 controls of UK Biobank resource followed by replication in 88,192 cases and 162,544 controls from CARDIoGRAMplusC4D.^17^ COVID-19 associated SNPs were obtained from a recent GWAS including 14,134 COVID-19 cases and 1,284,876 controls of European ancestry by the COVID-19 Host Genetics Initiative.^27^ We performed two-sample MR analyses^34^ using the TwoSampleMR R package.^39^ The instrumental variables (IVs) were independent *cis*-eQTL variants (LD r^2^ <0.1) from this study. The primary analysis used the inverse variance weighted (IVW) method. We also assessed heterogeneity of the IVs in each gene and conducted sensitivity analysis using the MR-Egger method to test for potential horizontal pleiotropy. We also performed the median-based method^40^ and mode-based method^41^ when heterogeneity was present in MR analyses due to outliers among the IVs^42^. We reported putative causal genes if Bonferroni correction *p* < 0.05/n (n is the number of genes tested).

### Replication analyses

A previous study reported 10,914 *cis-*eQTL variant-eGene pairs and 269 *trans* pairs (FDR < 0.05) through RNA-sequencing of 922 individuals.^21^ We performed replication analyses using the reported *cis- and trans-*eQTL variant-eGene pairs in conjunction with the pairs in the present study.^21^ We also used the *cis*-eQTL database generated from GTEx whole blood (version 8) (https://www.gtexportal.org/home/datasets) for replication of our *cis*-QTL findings. Whole genome sequencing and RNA-seq were conducted in whole blood of 755 samples in GTEx. The replication was only performed using the *cis*-eQTL-variant-eGene pairs generated by 8,372,247 SNPs and 20,188 gene transcripts that were found in common between our study and GTEx. Because this study was aimed to provide eQTL resource for the broad scientific community, we present replication results using both *p* < 5e-8 and *p* < 1e-4 for replicating *cis*-eQTL variant-eGene pairs.

## Data Availability

The data is available at dbGaP via TOPMed commons

https://topmed.nhlbi.nih.gov

https://www.ncbi.nlm.nih.gov/gap/

## Author contribution

C.L. wrote the main manuscript text; R.J., C.L., X.S., and P.J.M. performed statistics analyses and prepared the tables and figures; Y.W., G.M.P., and A.N.P. cleaned RNAseq data; N.L.H-C., and A.N.P. coordinate data acquisition; D.L., R.J., J.M., A.K., M.S., T.H., S-J.H., H.B., B.T., P.J.M., and R.S.V. reviewed/edited the manuscript; D.C.,M.F., and N.S. performed data sharing; R.S.V. and D.L. provided funding for whole genome sequencing and RNAseq.

---

**Supplemental Figure 1.**
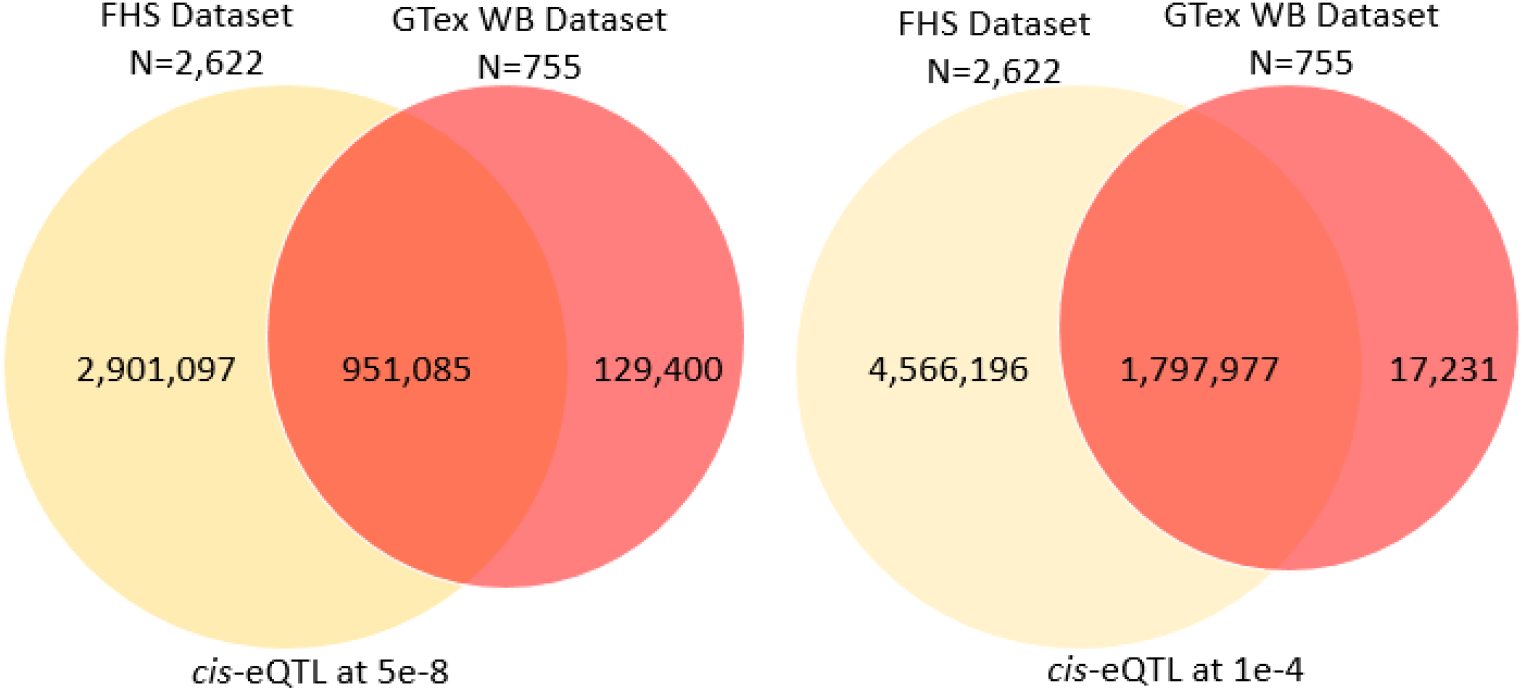

